# Investigating spatiotemporal patterns of the COVID-19 in São Paulo State, Brazil

**DOI:** 10.1101/2020.05.28.20115626

**Authors:** Enner Alcântara, José Mantovani, Luiz Rotta, Edward Park, Thanan Rodrigues, Fernando Campos Carvalho, Carlos Roberto Souza Filho

## Abstract

As of May 16th, 2020, the number of confirmed cases and deaths in Brazil due to COVID-19 hit 233,142 and 15,633, respectively, making the country one of the most affected by the pandemic. The State of Sao Paulo (SSP) hosts the largest number of confirmed cases in Brazil, with over 60,000 cases to date. Here we investigate the spatial distribution and spreading patterns of COVID-19 in the SSP by mapping the spatial autocorrelation and the clustering patterns of the virus in relation to the population density and the number of hospital beds. Moran’s I and LISA clustering analysis indicated that Sao Paulo City is a significant hotspot for both the confirmed cases and deaths, whereas other cities across the State could be considered colder spots. Bivariate Moran’s I also showed that the population density is a key indicator for the number of deaths, whereas the number of hospital beds is less related, implying that the fatality depends substantially on the actual patient’s condition. Social isolation measures throughout the SSP have been gradually increasing since early March, an action that helped to slow down the emergence of the new confirmed cases, highlighting the importance of the safe-distancing measures in mitigating the local transmission within and between cities in the SSP.

## 1. Introduction

In December 2019, health units in Wuhan City, Hubei Province, China, reported the occurrence of patients with pneumonia, of unknown cause, possibly epidemiologically associated with a seafood and live animals’ market. On December 31 of that year, the Chinese Center for Disease Control and Prevention (China CDC) sent a team to that province to accompany health authorities in conducting an epidemiological and etiological investigation (Zhu et al. 2019). The agent has been identified as a new coronavirus: SARS-COV-2. Simultaneously, the World Health Organization (WHO) was also informed. The first meeting of the Emergency Committee, convened by the WHO on January 23, 2020, abided to International Health Regulations (IHR). The possibility of the occurrence constituting a Public Health Emergency of International Importance (ESPII) was discussed (WHO, 2005). On a subsequent meeting on January 30, the ESPII status was consolidated (WHO, 2005), given the rapid escalation on the number of cases in China, as well as in other countries. The name of the new disease was assigned in February 2020, in reference to the type of virus and the year the epidemic started: Coronavirus disease – 2019 (COVID-19) (WHO, 2019). Until the end of February, the number of confirmed cases reached 80 thousand in China, totaling about 2,900 deaths. In other countries the number of infected people during the same period reached 6 thousand, totaling about 90 deaths.

In Brazil, the monitoring of cases began in January 2020. On January 22, the Emergency Operation Center (COE) of the Ministry of Health, coordinated by the Health Surveillance Secretariat, was activated to objectively plan and organize activities related to the control of local contamination, as well as international monitoring. A contingency plan was activated on January 27, and on February 3 the epidemic was declared a Public Health Emergency of National Importance (ESPIN) (Brazil, 2020). The first confirmed cases of COVID-19 in Brazil were reported around the end of February in 2020 and referred to male individuals, residing in the city of São Paulo, who had just returned from a trip to Italy. The cases were imported and until the beginning of March, evidence of local transmission of the virus was not found. The difficulty in containing the spread of the outbreak due to COVID-19 is that, although similar to the outbreaks caused by other coronaviruses, transmission can occur from asymptomatic cases; i.e., individuals who have not developed serious manifestations of the disease (Munster et al. 2020). Several studies have been carried out from the initial cases (Zhou et al. 2020; Lam et al. 2020; Lu et al. 2020; Wang et al. 2020) investigating several aspects associated with COVID-19, from the developmental process of this coronavirus to its clinical characteristics. According to the Brazilian Society of Infectious Diseases, in a statement dated March 12, 2020, the contagion capacity of SARS-COV-2 is 2.74; in comparative terms, the measles rate is 15.

Although the aggressive testing of cases nationwide and strong measures imposed by the government, such as social distancing, the number of both confirmed cases and deaths in Brazil grew exponentially throughout the country, reaching 233,142 and 15,633 (source: https://www.seade.gov.br/coronavirus/), respectively (fatality rate of 6.7%), as of May 17^th^ 2020.

Given this situation in Brazil, it is urgent to understand how COVID-19 is spreading in different regions of the country. In this paper, we investigated the spatial distribution patterns and dynamics of the COVID-19 in the State of São Paulo, the most affected State by the pandemic in Brazil.

### 1.1. Study area

The State of São Paulo State (SSP) is one of the 26 States of Brazil (IBGE, 2019, Figure 1). It hosts the largest population, with approximately 46 million citizens (> 20% of the country). As the wealthiest State, São Paulo is responsible for 33.9% of the Brazilian GDP and has the second-highest Human Development Index and GDP per capita. To date (May 16, 2020), among 233,142 confirmed cases in all regions of Brazil, most of them (41,830) are in the SSP (Brazil, 2020). The city of São Paulo, the State capital, hosts 26,273 of the confirmed cases and 2,135 deaths (SEADE, 2020).

**Figure 1:**
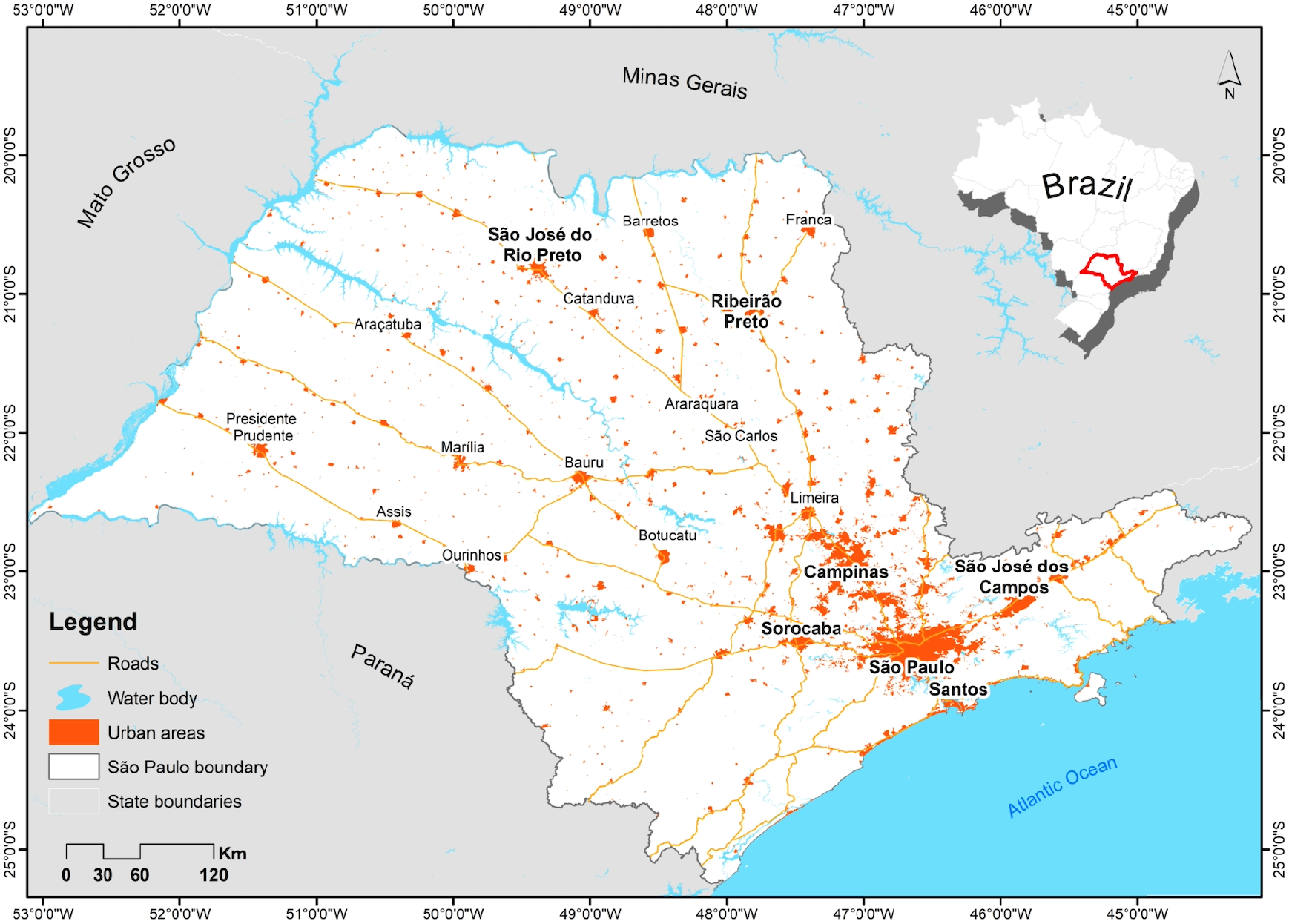
Map showing the location of the State of São Paulo in Brazil and main cities and roads.

## 2. Materials and Methods

### 2.1. Data sources and collection

The data about COVID-19, population density and social isolation index in São Paulo State was obtained from the São Paulo State Health Secretariat (https://www.seade.gov.br/coronavirus/) from March, 01 2020 to May, 08 2020. The social isolation index, which uses geolocation data provided by the nation’s main telecom firms and shows the percentage of the population that abiding by social isolation advice. The data about hospital beds were obtained from the DATASUS (https://datasus.saude.gov.br). The shape file of the Federal highways was downloaded from the Brazilian Institute of Geography and Statistics, IBGE (https://www.ibge.gov.br/pt/inicio.html), the official provider of geographic and statistical information about Brazil.

### 2.2. COVID-19 spatial association

Spatial autocorrelation statistics measure the degree to which a sub-region is similar to or different from its neighboring sub-regions for a particular indicator. Spatial autocorrelation may be evaluated globally as well as locally. Global measures summarize spatial autocorrelation, while local measures evaluate localized spatial autocorrelation within a study site. Moran’s I statistic is a measure of global spatial autocorrelation or clustering (Moran, 1950) and can be calculated as:

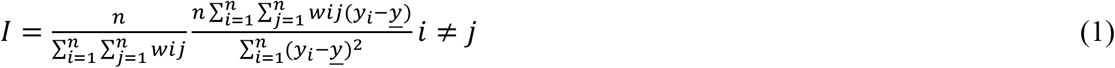

Where *n* is the number of spatial units indexed by *i* and *j*; *y_i_* and *y_j_* are the variables at points *i* and *j* (with *i* ≠ *j*); <u>y</u> is the mean of *y*; *wij* is an element of the weight matrix (*n* × *m*).

The Moran’s *I* range from –1 to 1. If *I* > 0, there is a positive spatial correlation; If *I* < 0, a negative spatial correlation exists. Statistical inference was based on 999 permutations testing the statistical significance (Anselin, 1995). We computed Moran’s *I* and both *z*-score and pseudo *p*-value to evaluate the statistical significance. In general, Moran’s *I* near +1 indicates clustering, while values near –1 indicates dispersion. The null hypothesis states that there is no spatial clustering of the values associated with the geographical units in the study site. The null hypothesis can be rejected when the pseudo *p*-value is small, and the *z*-score is large enough that it falls outside of the desired confidence level.

However, spatial correlation patterns may differ locally within the study site; therefore, measures of global spatial autocorrelation do not reflect the local spatial correlations within geographic units once there is a spatial heterogeneity. To overcome this, we used Anselin’s LISA (Local Indicators of Spatial Association), a measure to describe the heterogeneity of spatial association across different geographic units (Anselin, 1995). Anselin local Moran’s *I* can be defined as:

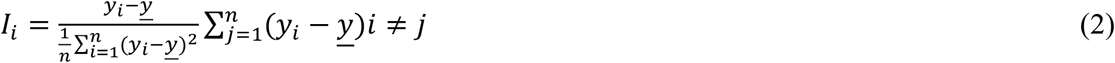

Where *j* is within distance of *i, I_i_* refers to the value of local Moran’s *I* at points i. The parameters *y*_i_, *y*_j_, <u>y</u> and *wij* have the same meaning as they are in equation 1.

The spatial autocorrelations between COVID-19 and deaths confirmed cases, hospital beds availability, human development index by cities and population density were tested using the bivariate Moran’s *I* statistics. The global Moran’s I describe the overall spatial relationship across all geographic units for the entire study site and can be calculated as:

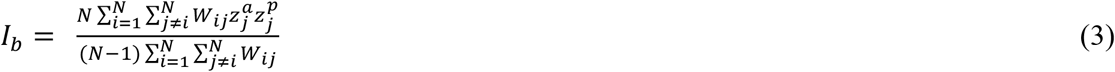

*I_b_* is the bivariate global spatial correlation index, *N* is the total number of geographic units, *W_ij_* is the spatial weight matrix, 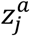 is the standardized COVID-19 cases or deaths, and 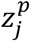 is standardization of hospital beds availability, human development index by cities and population density.

## 3. Results

### 3.1. COVID-19 confirmed cases spatial distribution

The first Brazilian COVID-19 confirmed case was reported on February 26, 2020, in the city of São Paulo. Shortly after and on March 11, 2020, the World Health Organization (WHO) declared that the COVID-19 as a pandemic. Figure 2 shows the COVID-19 confirmed cases from March, 01 2020 to May, 08 2020 had spread throughout the SSP. On March, 01 2020, SSP had 1 case and after 24 days the cases jumped to 722. Although the SSP Government declared partial lockdown on March, 24 2020, the confirmed cases reached 24,273 contaminated peoples by May, 08 2020.

**Figure 2:**
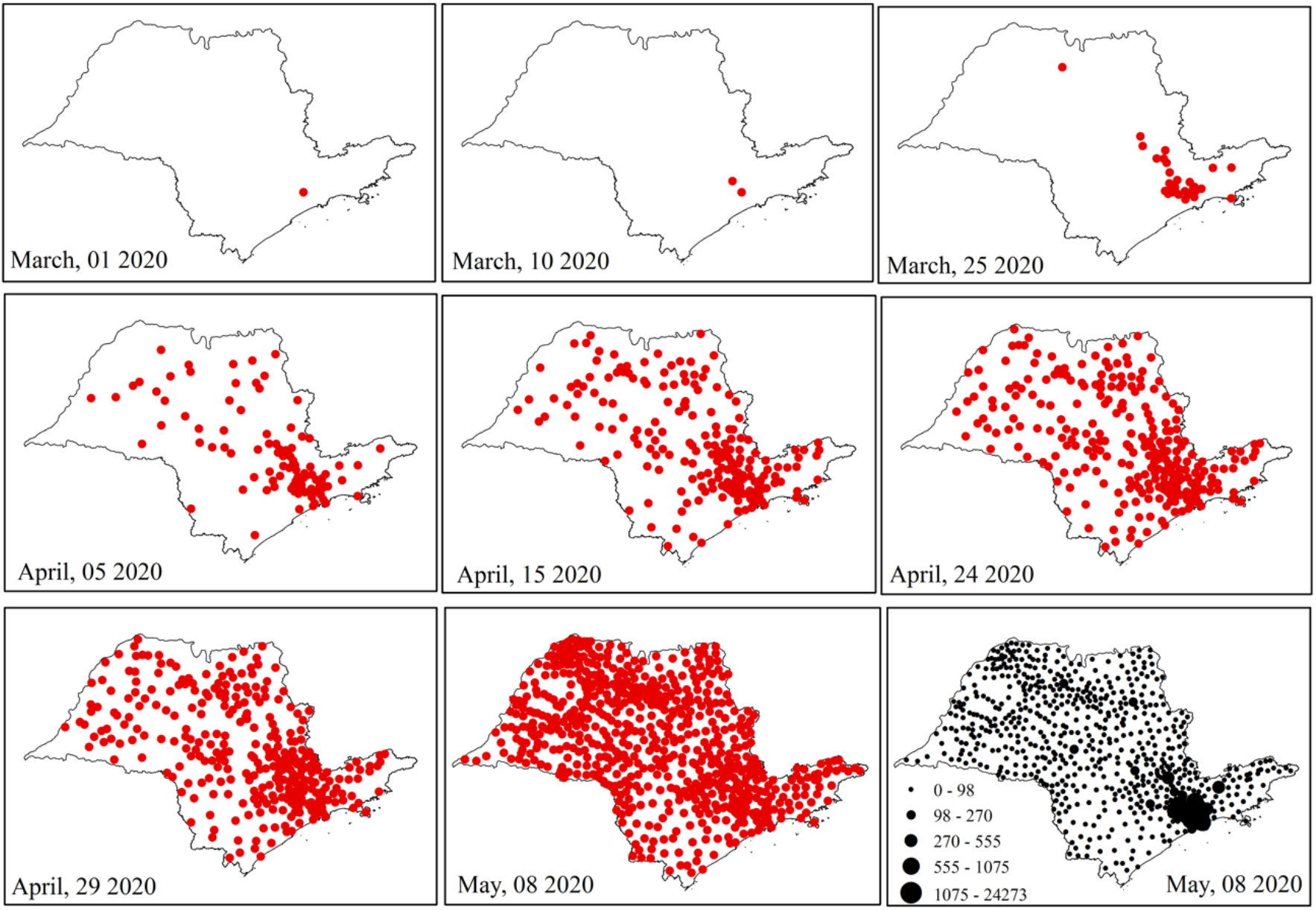
Spatial distribution of COVID-19 confirmed cases in São Paulo State.

The highest numbers of confirmed cases were registered in the São Paulo Metropolitan Area (SPMA), and the lowest for the littoral cities. Regarding the São Paulo coast, most cases coming from cities were shielded from the presence of tourists, who typically frequent the beaches on holidays and weekends. At least seven cities have taken measures to prevent the interplay of tourists and allowing only the movement of residents. This restriction of movement only allowed people working in essential services, such as medical or transportation, to enter the SPMA. The other cities along the coast maintained the policy of guiding the need for social isolation, the limiting of hotel services and the prohibiting loitering on the beaches. According to the state authority, if the number of cases on the coast and inland continues to grow in the same way as in the state capital, the entire state health system will collapse. Due to the highly heterogeneously distributed COVID-19 cases observed in the State, in the next section, the Moran’s I method is applied to map the level of clustering of the cases around the cities that are socio-economically intimately connected.

### 3.2. COVID-19 spatial autocorrelation

The calculated Moran’s *I* values are positive (0.062 and 0.066) for both confirmed cases and deaths, indicating the positive spatial autocorrelation. This indicates that there are similarities between values of confirmed cases and deaths with the spatial location (Figure 3).

**Figure 3:**
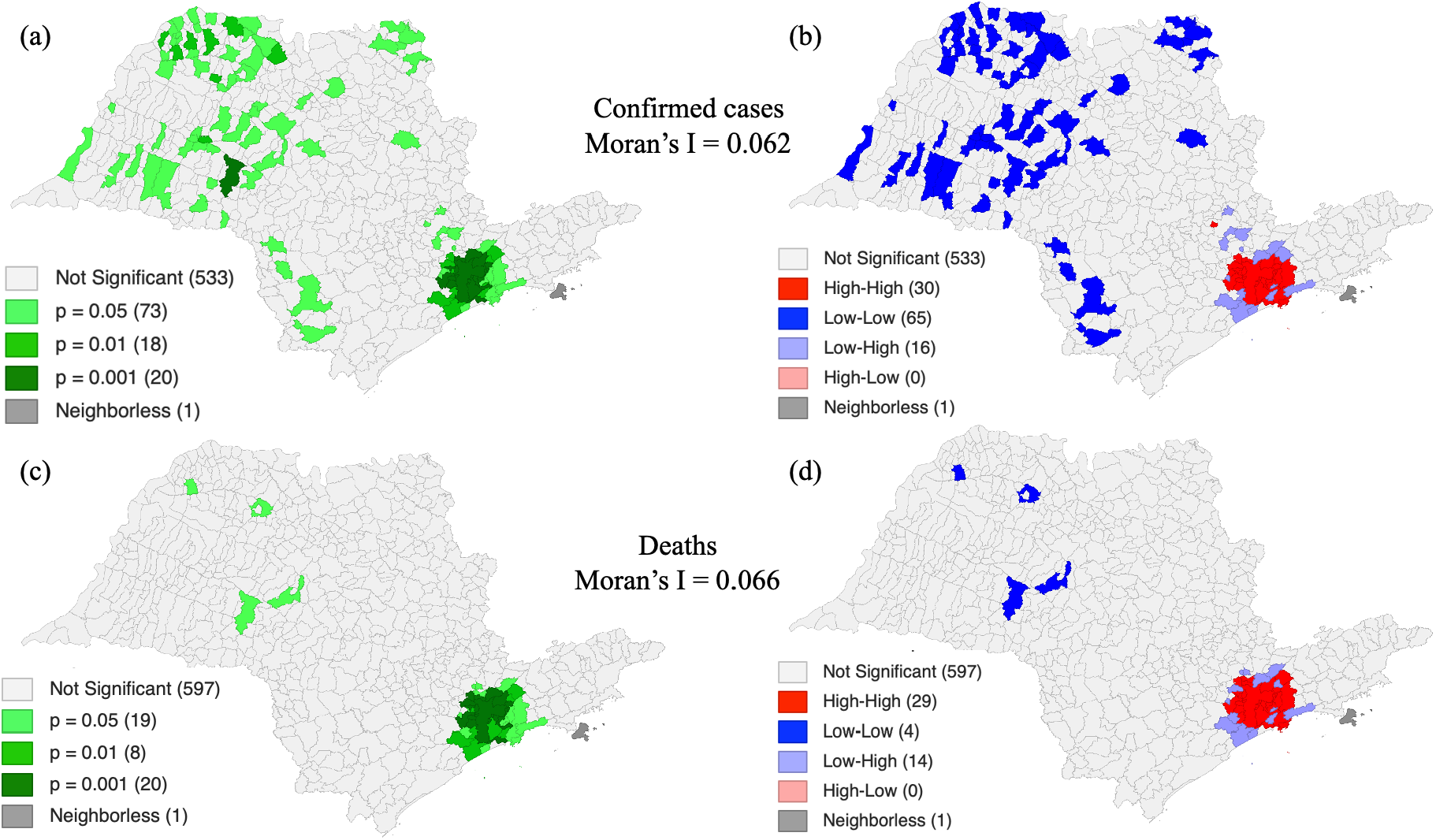
LISA’s significant maps for (a) confirmed cases and (c) deaths and LISA’s clusters maps for (b) confirmed cases and (d) deaths. LISA’s maps were based on data collected until May, 08 2020. The numbers in parentheses represent the number of cities. The total number of cities in São Paulo State is 645.

In Figures 3a and c, the LISA significant maps show the regions that have passed the local Moran’s significance test for both confirmed and deaths cases in São Paulo State. Based on that, Figure 3b highlights the main clusters of confirmed cases in the State, and the High-High class (red) means that these regions contain a high number of confirmed cases, relative to the average, surrounded by regions that also present a high number of cases. This class is represented by the metropolitan region of São Paulo, the most populous region of the State and where the first cases were reported.

On the other hand, the Low-Low class (blue) refers to a spatial association group whose regions show a low number of confirmed cases, i.e. below the average, surrounded by regions that also have low values. These regions are represented by small cities compared to the state capital and the places where the cases took longer to be reported (see the spatial distribution in Figure 2). Regarding the Low-High class (purple), regions with low confirmed cases are surrounded by regions with high confirmed cases. Although there are confirmed cases in the western region of the state, the death records are concentrated in the metropolitan region of São Paulo, where the number of confirmed cases is also high (Figure 3d).

### 3.3. Relationship between COVID-19 deaths and population density and hospital beds

The bivariate Moran’s *I* result show that a positive relationship between deaths and population density, i.e. 0.769, indicating that the population density is a key parameter to explain the number of deaths. On the other hand, the number of hospital beds yielded *I* = 0.070, implying that there is no relation with the number of deaths (Figure 4).

**Figure 4:**
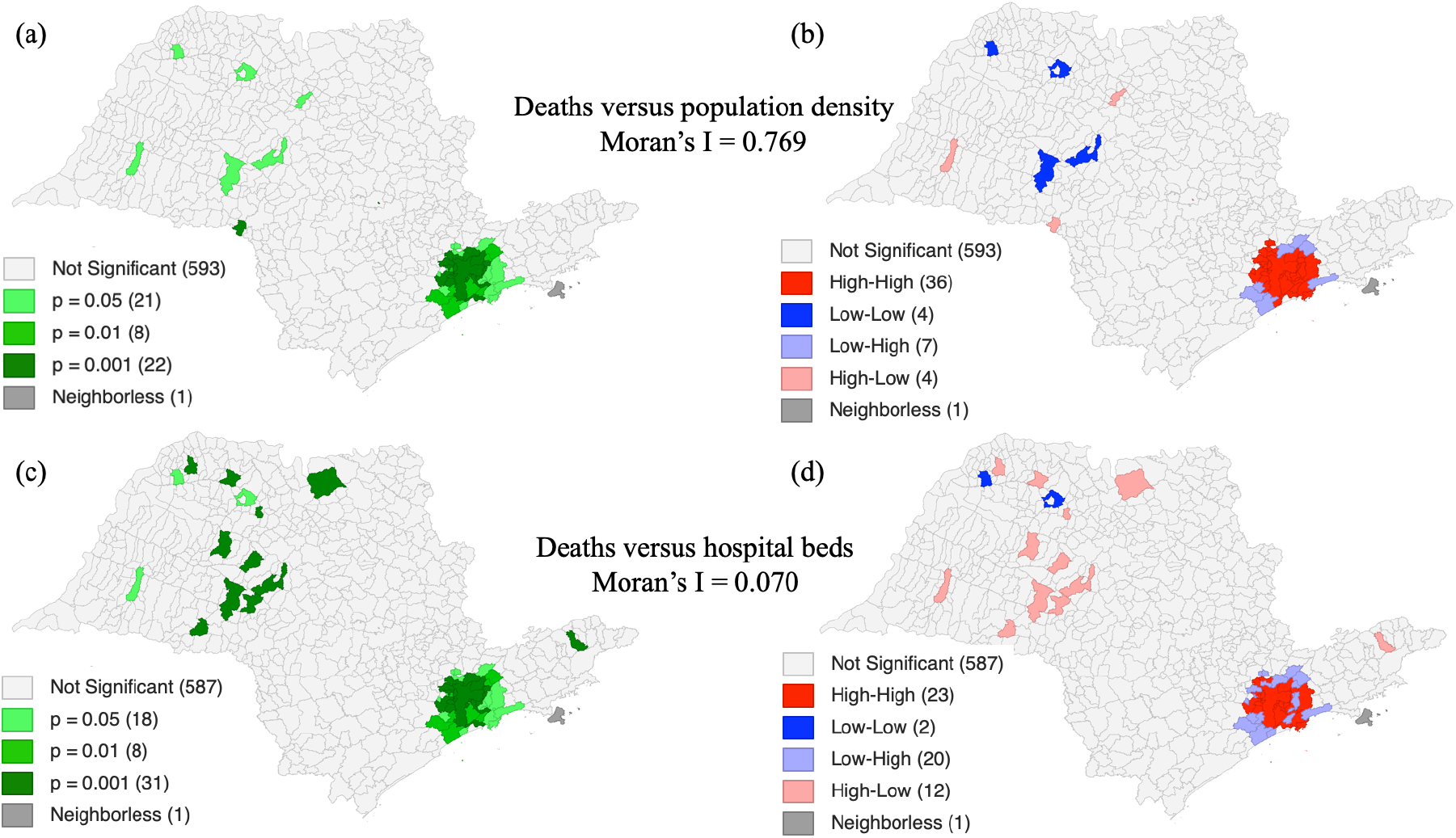
LISA’s significant maps for the correlation between deaths and population density(a) and between deaths and availability of hospital beds (c), and LISA’s clusters maps for the correlation between deaths and population density (b) and between deaths and availability of hospital beds (d). LISA’s maps are based on data collected until May, 08 2020, for the State of São Paulo.

The relationship between the number of deaths versus population density (Figure 4b) and deaths versus hospital beds (Figure 4d) shows a high-high cluster (mainly in the SPMA), surrounded by a low-low spatial association. The high-high means that this city has a high number of deaths and high population density, as well as a large number of hospital beds available (Figure 5). The low-low means the opposite. Cities classified as high-low and low-high indicate, respectively, that they had an increase in the death scenario with low population density and a small number of hospital beds, and a smaller death number with a high number of hospital beds. In this particular cluster, the city is in the transition between spatial regimes (e.g. moving from low contamination regime to a high contamination rate or vice versa). That is, these cities are moving to higher death rates or the hospital infrastructure and the social isolation is working.

**Figure 5:**
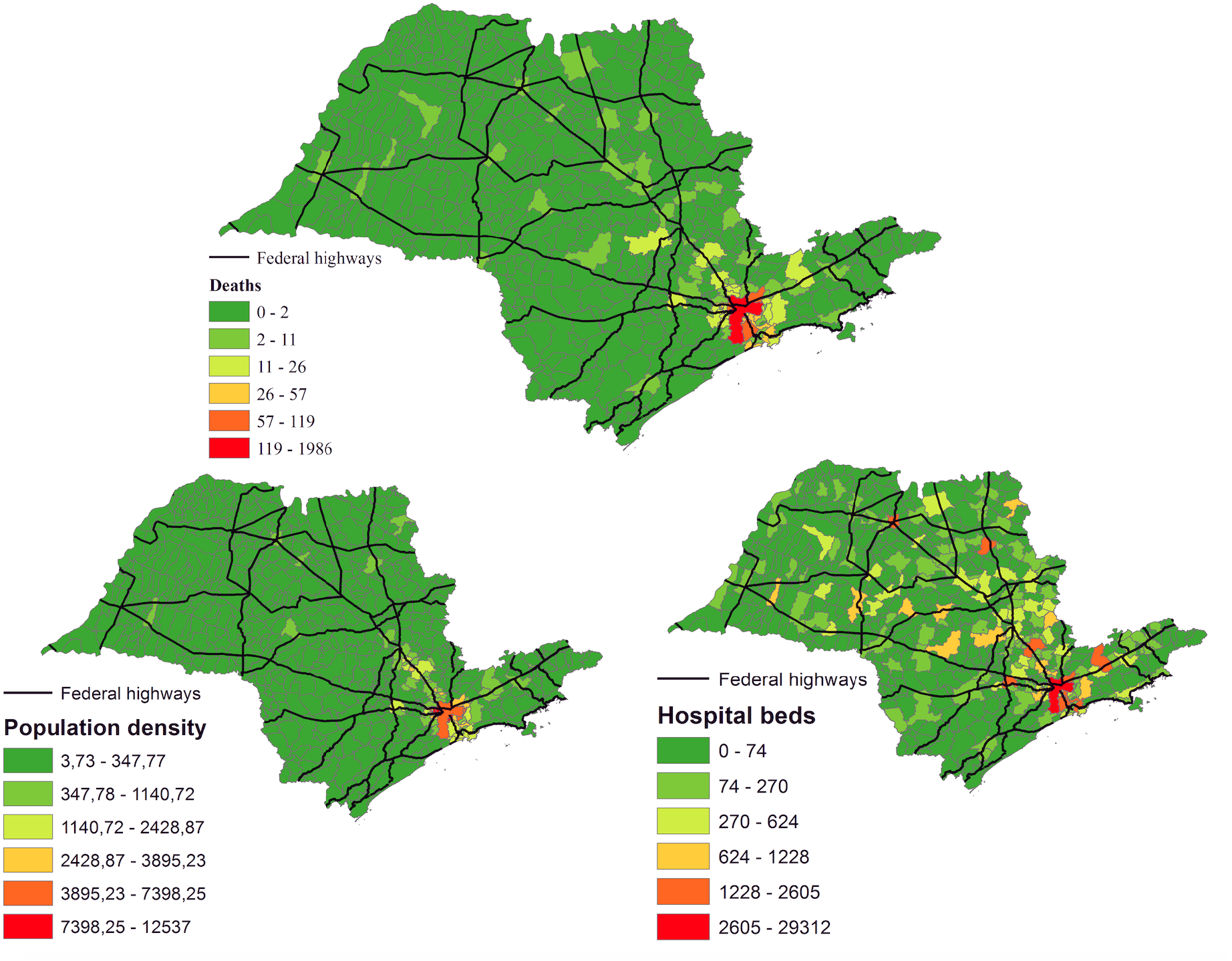
Deaths by COVID-19, population density and hospital beds in the State of São Paulo.

### 3.4. Evolution of social isolation

The social isolation maps in the cities computed with data collected on March, 05 and 12 2020, show that the majority reached an isolation fraction of less or up to 40%. This pattern only varied slightly until March, 17 2020, where the isolation started to increase up to 50% for some regions (Figure 6).

**Figure 6:**
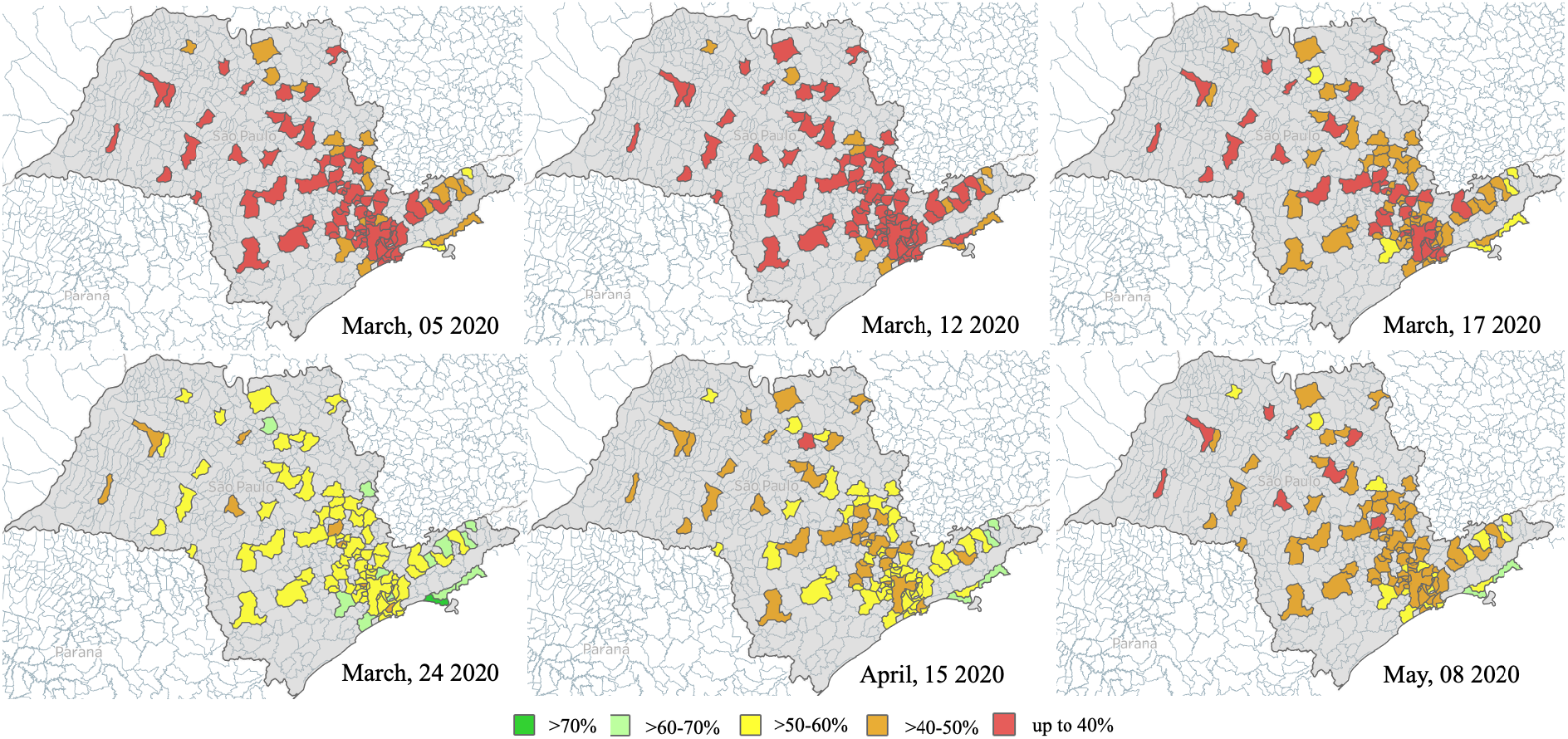
Evolution of social isolation in State of São Paulo State. Source: Adapted from São Paulo (2020).

On April, 15 2020 and May, 08 2020 social isolation maps showed that the cities reduced the isolation from 50–60% to 40–50%. According to São Paulo (2020) since the partial lockdown of the state started on March, 24 2020, social isolation varied from 54% (March 24), achieving a minimum of 47% (April 09) and a maximum of 59% (several dates), with an average of 54%.

## 4. Discussion

The results on spatial distribution patterns of the COVID-19 cases in the Sao Paulo State showed a clear aggregation pattern among cities: high rate of contamination surrounded by the cities with similar level of contamination and cities with low contamination surrounded by the cities with low values. The highest contaminated cluster is on the SPMA and the lower is in the countryside. The same pattern was observed for the death case map, but the low-low cases were restricted only to four cities in the countryside, Marília (population = 232,006), Monte Aprazível (population = 24,794), Pirajuí (population = 24,546) and Palmeira D’oeste (population = 9,584). These same cities are aggregated as low-low for the relationship between deaths and population density and high-low for the relationship between deaths and available hospital beds.

Auler et al. (2020) and Prata et al. (2020) modeled the influence of temperature and humidity in the COVID-19 cases in Brazil and found that cities with lower temperatures tended to have more cases. Their findings are in line with our results, since SPMA – which was a higher number of cases – has lower temperature, and the countryside cities with higher temperature showed lower cases. However, temperature and humidity are not sufficient for modelling the COVID-19 spreading patterns in São Paulo State, due to the complexity of human factors such as the massive daily movement of people.

The social isolation maps showed that for the most populated cities, soon after the partial lockdown, the isolation increased but days later the isolation rates decreased. Nakada et al. (2020) showed that due to the partial lockdown, the air quality improved due to the reduction of cars in the SSP, which corroborates with our results. The same pattern was observed for cities in the countryside.

The lack of spatial autocorrelation between deaths and hospital beds highlights that its quantity per se is not directly related to the chances of a patient’s survival. Once hospital beds are not equipped with a ventilator and an experienced health care team, critical patients might die. Requia et al. (2020) mapped the risk of the Brazilian health care system and highlighted that the presence of ventilators can decrease the fatality rate and therefore needed to be considered as a key parameter to analyze the fatality rate due to the COVID-19.

Figure 5 reveals that São Paulo city, the most populated city in the State, had the highest rate of contamination and deaths. In contrast, the countryside might be protected from the virus if the state government regulates movement around the State. The importance of the lockdown in curbing the transmission of COVID-19 in the state is already shown, as well as in other countries (Acter et al. 2020). It becomes even more important given that the first case in most of the cities were imported (Candido et al. 2020). Social isolation is essential in the initial stage of the spread of the virus, limiting contamination to a certain area (Kraemer et al., 2020). Therefore, it is timely that we develop sustainable safe-distancing measures to mitigate the local transmission within and between cities.

## 5. Conclusion

With a number of confirmed cases and deaths of 233,142 and 15,633, respectively, as of May 16^th^, 2020, Brazil currently ranks among the most affected countries by the COVID-19. The SSP contains the largest number of confirmed cases in Brazil, with over 60,000 cases to date. In this work, we investigated the spatial distribution and spread patterns of the COVID-19 in the SSP by mapping the spatial autocorrelation and the clustering patterns in relation to the population density and the number of hospital beds. The LISA analysis indicated that Sao Paulo city is a significant hotspot for both the confirmed and death cases, while other cities across the state could be considered cold spots. Bivariate Moran’s I also showed that the population density is a key indicator for the number of deaths while the number of hospital beds is less so, implying that the fatality depends substantially on the actual patient’s condition. Social isolation measures throughout the State has been gradually increasing since the early March that helped to slow down the emergence of the new confirmed cases, highlighting the importance of the safe-distancing measures in mitigating the local transmission within and between cities in the state.

## Data Availability

All data used are free and can be downloaded directly from: https://www.seade.gov.br/coronavirus/

## Acknowledgements

E.A. and C.R.S.F. acknowledge the Brazilian National Council for Scientific and Technological Development (CNPq) for research grants 303169/2018–4 and 309712/2017–3, respectively.

## References

Acter, T., Uddin, N., Das, J., Akhter, A., Choudhury, T.R., Kim, S. Evolution of severe acute respiratory syndrome coronavirus 2 (SARS-CoV-2) as coronavirus disease 2019 (COVID-19) pandemic: A global health emergency. Science of The Total Environment. 730 (2020). Article 138996.

Anselin L. Local indicators of spatial association–LISA. Geographical Analysis. 27 (1995). pp. 93–115.

Auler, A.C., Cássaro, F.A.M., Silva, V.O., Pires, L.F. Evidence that high temperatures and intermediate relative humidity might favor the spread of COVID-19 in tropical climate: a case study for the most affected Brazilian cities. Science of The Total Environment. 2020. 729. Article 139090.

Brazil. Ministry of Health – COVID19 – Coronavirus Panel. https://covid.saude.gov.br/. (2020), Accessed 14th May 2020.

Candido, D.D.S. et al. Routes for COVID-19 importation in Brazil. Journal of Travel Medicine, taaa042, (2020). https://doi.org/10.1093/jtm/taaa042.

IBGE (Brazilian Institute of Geography and Statistics), Population Estimation 2019. (2019). (Brasília).

Kraemer et al. The effect of human mobility and control measures on the COVID-19 epidemic in China. Science 368 (2020). pp. 493–497.

Lam, T.T.Y., Shum, M.H.H., Zhu, H.C., Tong, Y.G., Ni, X.B., Liao, Y.S., Wei, W., Cheung, W.Y.M., Li, W.J., Li, L.F., Leung, G.M., Holmes, E.C., Hu, Y.L., Guan, Y. Identification of 2019-nCoV related coronaviruses in Malayan pangolins in southern China. BioRxiv. (2020). https://doi.org/10.1101/2020.02.13.945485.

Lu, R., et al. Genomic characterization and epidemiology of 2019 novel coronavirus: implications for virus origins and receptor binding. Lancet. (2020).https://doi.org/10.1016/S0140-6736(20)30251-8.

Moran, P.A.P. Notes on continuous stochastic phenomena. Biometrika. 37. (1950). pp. 17– 23.

Munster, V.J., Koopmans, M., Van Doremalen N., Van Riel, D., Wit, E. A novel coronavirus emerging in china – key questions for impact assessment. New England Journal of Medicine. 382 (2020). pp. 692–4.

Nakada, L.Y.K., Urban, R.C. COVID-19 pandemic: Impacts on the air quality during the partial lockdown in São Paulo state, Brazil. Science of the Total Environment. 730. (2020). Article 139087.

Prata, D.N., Rodrigues, W., Bermejo, P.H. Temperature significantly changes COVID-19 transmission in (sub)tropical cities of Brazil. Science of The Total Environment. 729. (2020). Article 138826.

Requia, W.J., Kondo, E.K., Adams, M.D., Gold, D.R., Struchiner, C.J. Risk of the Brazilian health care system over 5572 municipalities to exceed health care capacity due to the 2019 novel coronavirus (COVID-19). Science of the Total Environment. 730. (2020). Article 139144

São Paulo. São Paulo State – Social Isolation Intelligent Monitoring System. https://www.saopaulo.sp.gov.br/coronavirus/isolamento/ (2020), Accessed 14 May 2020. SEADE. Coronavirus. https://www.seade.gov.br/coronavirus/ (2020), Accessed 14th May 2020.

Wang, D., Hu, B., Hu, C., Zhu, F., Liu, X., Zhang, J., Wang, B., Xiang, H., Cheng, Z., Xiong, Y., Zhao, Y., Li, Y., Wang, X., Peng, Z. Clinical characteristics of 138 hospitalized patients with 2019, novel coronavirus–infected pneumonia in Wuhan China. JAMA. 323. (2020). pp. 1061–1069. https://doi.org/10.1001/jama.2020.1585.

WHO – World Health Organization. Statement on the meeting of the International Health Regulations (2005) Emergency Committee regarding the outbreak of novel coronavirus (2019-nCoV) [Internet]. Geneva: World Health Organization; 2020 [cited 2020 Mar 4]. Available https://www.who.int/news-room/detail/23-01-2020-statement-on-the-meeting-of-the-international-health-regulations-(2005)-emergency-committee-regarding-the-outbreak-of-novel-coronavirus-(2019-ncov).

WHO – World Health Organization. Novel coronavirus (2019-nCoV): situation report – 22 [Internet]. Geneva: World Health Organization; 2020 [cited 2020 Mar 4]. Available from:https://www.who.int/docs/default-source/coronaviruse/situation-reports/20200211-sitrep-22-ncov.pdf?sfvrsn=fb6d49b1_2.

Zhu, N. et al. A novel coronavirus from patients with pneumonia in China. New England Journal of Medicine. 382 (2020). pp. 727–733.

Zhou P. A pneumonia outbreak associated with a new coronavirus of probable bat origin. Nature. 579. (2020). pp. 270-273. https://doi.org/10.1038/s41586-020-2012-7".

